# Methodological Rigor in COVID-19 Clinical Research – A Systematic Review and Case-Control Analysis

**DOI:** 10.1101/2020.07.02.20145102

**Authors:** Richard G. Jung, Pietro Di Santo, Cole Clifford, Graeme Prosperi-Porta, Stephanie Skanes, Annie Hung, Simon Parlow, Sarah Visintini, F. Daniel Ramirez, Trevor Simard, Benjamin Hibbert

## Abstract

**Objective:** To systematically evaluate the quality of reporting of currently available COVID-19 studies compared to historical controls.

**Design:** A systematic review and case-control analysis

**Data sources:** MEDLINE, Embase, and Cochrane Central Register of Controlled Trials until May 14, 2020

**Study selection:** All original clinical literature evaluating COVID-19 or SARS-CoV2 were identified and 1:1 historical control of the same study type in the same published journal was matched from the previous year

**Data extraction:** Two independent reviewers screened titles, abstracts, and full-texts and independently assessed methodological quality using Cochrane Risk of Bias Tool, Newcastle-Ottawa Scale, QUADAS-2 Score, or case series checklist.

**Results:** 9895 titles and abstracts were screened and 686 COVID-19 articles were included in the final analysis in which 380 (55.4%) were case series, 199 (29.0%) were cohort, 63 (9.2%) were diagnostic, 38 (5.5%) were case-control, and 6 (0.9%) were randomized controlled trials. Overall, high quality/low-bias studies represented less than half of COVID-19 articles - 49.0% of case series, 43.9% of cohort, 31.6% of case-control, and 6.4% of diagnostic studies. We matched 539 control articles to COVID-19 articles from the same journal in the previous year for a final analysis of 1078 articles. The median time to acceptance was 13.0 (IQR, 5.0-25.0) days in COVID-19 articles vs. 110.0 (IQR, 71.0-156.0) days in control articles (p<0.0001). Overall, methodological quality was lower in COVID-19 articles with 220 COVID-19 articles of high quality (41.0%) vs. 392 control articles (73.3%, p<0.0001) with similar results when stratified by study design. In both unadjusted and adjusted logistic regression, COVID-19 articles were associated with lower methodological quality (odds ratio, 0.25; 95% CI, 0.20 to 0.33, p<0.0001).

**Conclusion:** Currently published COVID-19 studies were accepted more quickly and were found to be of lower methodological quality than comparative studies published in the same journal. Given the implications of these studies to medical decision making and government policy, greater effort to appropriately weigh the existing evidence in the context of emerging high-quality research is needed.

**Study registration:** PROSPERO: CRD42020187318

## Introduction

The severe acute respiratory syndrome coronavirus 2 (**SARS-CoV-2**) pandemic spread globally in early 2020 with major health and economic consequences. This was shortly followed by an exponential increase in scientific publications related to the coronavirus disease 2019 (**COVID- 19**) in order to rapidly elucidate the natural history and identify diagnostic and therapeutic tools.^1^

While a need to rapidly disseminate information to the medical community, governmental agencies, and general public was paramount - major concerns have been raised regarding scientific rigor in the currently published literature.^2^ Poorly conducted studies may originate from failure at any of the four consecutive stages: 1) choice of research question relevant to patient care, 2) quality of research design, 3) adequacy of publication, and 4) quality of research reports.^3^ Furthermore, evidence-based medicine fundamentally relies on the hierarchy of medical evidence ranging from the highest level of randomized controlled trials (**RCT**) to the lowest level of case series in order to inform medical practice and generate clinical practice guidelines.^4^

Given the implications for clinical medicine, policy decision making, and the widely expressed concern of methodological rigor and rapidity of publication,^5^ we sought to perform a formal evaluation of the quality of COVID-19 literature. Herein, we performed a systematic review to identify COVID-19 clinical literature and generated a historical control in order to formally evaluate the following: 1) quality of COVID-19 literature using established quality scores, 2) quality of COVID-19 literature stratified by median time to acceptance, geographical regions, and impact factor, and 3) comparison of COVID-19 literature to the historical control in order to evaluate differences in study quality.

## Methods

A systematic literature search was conducted on May 14, 2020 (**PROSPERO: CRD42020187318**) and conducted according to the Preferred Reporting Items for Systematic Reviews and Meta-Analyses guidelines.

### Data sources and searches

The search was created in MEDLINE using a combination of key terms and index headings related to COVID-19 and translated to the remaining bibliographic databases (**Supplementary Table 1**). The searches were conducted in MEDLINE (Ovid MEDLINE(R) ALL 1946-), Embase (Ovid Embase Classic + Embase 1947-) and the Cochrane Central Register of Controlled Trials (from inception). Search results were limited to English-only publications, and a publication date limit of January 1, 2019 to present was applied. In addition, a Canadian Agency for Drugs and Technologies in Health search filter was applied in MEDLINE and Embase to remove animal studies, and comment, newspaper article, editorial, letter and note publication types were also eliminated. Search results were exported to Covidence and duplicates were eliminated using the platform’s duplicate identification feature.

### Study selection and quality assessment

We included all clinical studies from case series, observational studies, diagnostic studies, and RCTs evaluating COVID-19. For diagnostic studies, the reference standard was considered a nasopharyngeal swab followed by reverse transcriptase polymerase chain reaction in order to detect SARS-CoV-2. We excluded studies which were exploratory or pre-clinical in nature (ie. *in vitro* or animal studies), case reports, case series <5 patients, studies published in a language other than English, reviews, methods or protocols, and other coronavirus variants such as the Middle East Respiratory Syndrome.

Title and abstracts were evaluated by two independent reviewers using Covidence (Melbourne, Australia) and all discrepancies were resolved by consensus. Articles that were selected for full review were independently evaluated by two reviewers for quality assessment. A historical comparator group was generated by identifying reports of the same study design matched in a 1:1 fashion. These were identified by searching the same journal starting in the edition 12 months prior to publication and proceeding forward in a temporal fashion until the first matched study was identified. Quality assessment was similarly conducted on the identified articles. If no comparator manuscript was found, the corresponding COVID-19 article was excluded from the comparative analysis.

### Statistical analysis

Continuous variables were reported as mean ± SD or median ± IQR as appropriate, and categorical variables were reported as proportions (%). Normally distributed continuous variables were compared using Mann-Whitney U-test and categorical variables and quality scores were compared by Chi-squares, Fisher’s exact test, or Kruskal-Wallis test.

The primary outcome of interest was to evaluate the quality of COVID-19 by study type using Newcastle-Ottawa Scale (**NOS**) for case-control and cohort studies, QUADAS-2 tool for diagnostic studies,^6^ Cochrane Risk of Bias for RCTs,^7^ and a score derived by Murad et al. for case series.^8^ Prespecified secondary outcomes were comparison of quality scores by: i) median time to acceptance, ii) impact factor, iii) geographical region, and iv) historical comparator. Good quality of NOS was defined as 3+ on selection and 1+ on comparability and 2+ on outcome/exposure domains. High quality case series was defined as a score ≥3.5. Time to acceptance was defined as the time between submission to acceptance which captures peer review and editorial decisions. Geographical region was stratified on a continent basis into Asia/Oceania, Europe/Africa, and Americas (North and South America).

The association of high methodological quality with COVID-19 and control studies, median time to publication, high journal impact factor, and geographical region was assessed by simple and multivariable logistic regression and was reported as odds ratio (**OR**) with 95% confidence intervals. All statistical analyses were performed using SAS v9.4 (SAS Institute, Inc., Cary, NC, USA). Statistical significance was defined as P < 0.05. All figures were generated using GraphPad Prism v8 (GraphPad Software, La Jolla, CA, USA).

### Patient and public involvement

No patients were involved in generating the research question and outcome, nor were they involved in the design, conduct, reporting, or dissemination plans for our research.

## Results

### Article selection

A total of 14,787 COVID-19 papers were identified as of May 14, 2020 and 4892 duplicates were removed. 9895 titles and abstracts were screened, and 794 full texts were reviewed for eligibility. Over 108 articles were excluded for improper study design, patient population, non-English manuscript, duplicates, wrong outcomes, and published in a non-peer review journal. Finally, 686 articles were identified and underwent quality assessment (**Figure 1**).

**Figure 1.**
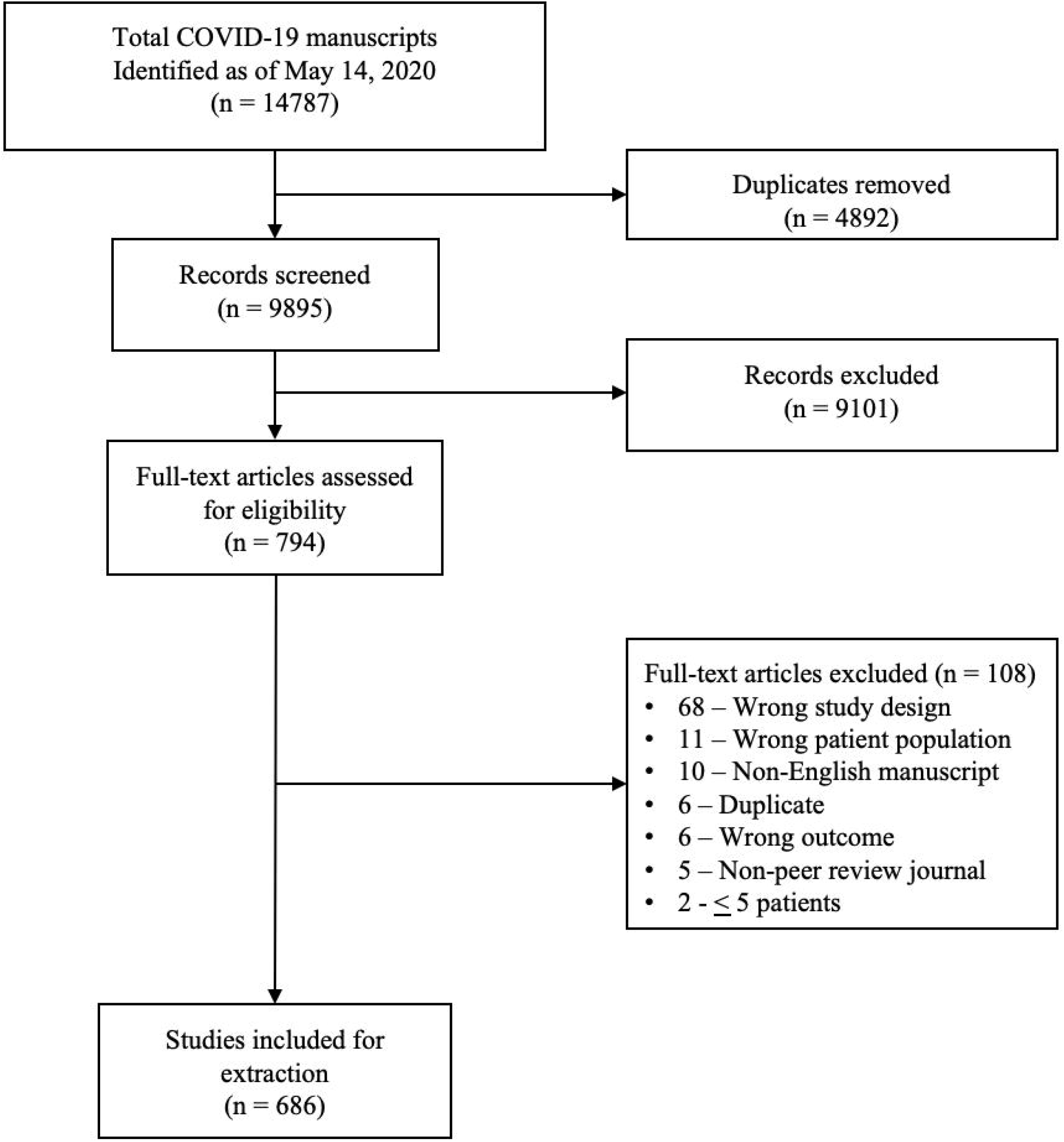
Literature search and selection of COVID-19 articles.

### COVID-19 literature quality

The majority of studies originated in Asia/Oceania with 486 (70.9%) followed by Europe with 122 (17.8%) and the Americas with 78 (11.4%). Of included studies, 380 (55.4%) were case series, 199 (29.0%) were cohort, 63 (9.2%) were diagnostic, 38 (5.5%) were case-control, and 6 (0.9%) were RCTs. Most studies (86.0%) were retrospective in nature, a total of 620 (90.4%) reported the sex of the cohort, and 7 (1.0%) studies determined their sample size *a priori*. The method of SARS-CoV-2 diagnosis was reported in 558 studies (81.3%) and ethics approval was received in 556 studies (81.0%). Finally, the median impact factor of identified manuscripts was 4.7 (IQR, 2.9-7.6) and median time to acceptance was 13.0 (IQR, 5.0-25.0) days (**Table 1**).

**Table 1.** Characteristics of COVID-19 Clinical Literature Until May 14, 2020.

|  | COVID-19 articles (n=686) |  | Matched articles (n=1078) |  |  |  | P-value |
| --- | --- | --- | --- | --- | --- | --- | --- |
|  |  |  | COVID-19 (n=539) |  | Control (n=539) |  |  |
|  | N | % | N | % | N | % |  |
| Geographic Region |  |  |  |  |  |  | <0.0001 |
| Asia/Oceania | 486 | 70.9 | 377 | 69.9 | 167 | 31 |  |
| Europe/Africa | 122 | 17.8 | 99 | 18.4 | 176 | 32.7 |  |
| Americas | 78 | 11.4 | 63 | 11.7 | 196 | 36.4 |  |
| Type of Article |  |  |  |  |  |  | 0.91 |
| Original Research | 614 | 89.5 | 486 | 90.2 | 484 | 89.8 |  |
| Letter | 35 | 5.1 | 26 | 4.8 | 29 | 5.4 |  |
| Communication | 37 | 5.4 | 27 | 5 | 26 | 4.8 |  |
| Retrospective study | 590 | 86.0 | 459 | 86.2 | 386 | 71.6 | <0.0001 |
| Sex reported | 620 | 90.4 | 484 | 89.8 | 483 | 89.6 | 0.92 |
| Sample size calculated | 7 | 1.0 | 4 | 0.7 | 34 | 6.3 | <0.0001 |
| Method of SARS-CoV-2 Diagnosis |  |  |  |  |  |  |  |
| PCR | 548 | 79.9 | 437 | 81.1 | N/A | N/A |  |
| ELISA | 2 | 0.3 | 1 | 0.2 | N/A | N/A |  |
| Physical exam only | 5 | 0.7 | 4 | 0.7 | N/A | N/A |  |
| Multiple | 3 | 0.4 | 2 | 0.4 | N/A | N/A |  |
| Unknown | 128 | 18.7 | 95 | 17.6 | N/A | N/A |  |
| Ethics approval received | 556 | 81.0 | 433 | 80.3 | 451 | 83.7 | 0.48 |
| Impact Factor - median (IQR) | 4.7 | (2.9-7.6) | 4.7 | (2.9-7.6) | 4.7 | (2.9-7.6) |  |
| Time to Acceptance - median (IQR) | 13.0 | (5.0-25.0) | 13.0 | (5.0-25.0) | 110.0 | (71.0-156.0) | <0.0001 |

Overall, good quality/low-bias studies represented less than half of identified COVID-19 articles - 49.0% of case series with a mean score (out of 5) (±SD) of 3.3 ± 1.1, 43.9% of cohort with a mean score (out of 8) of 5.8 ± 1.5, 31.6% of case-control with a mean score (out of 8) of 5.5 ± 1.9, and low bias present in 4 (6.4%) diagnostic studies (**Figure 2A**). In prespecified secondary analysis, rapid time from submission to acceptance (34.4% vs. 46.3%, p=0.01, **Figure 2B**) and low impact factor journals (<10) resulted in lower study quality (38.8% vs 68.0%, p<0.0001, **Figure 2C**). Finally, studies originating in either Americas or Asia/Oceania demonstrated higher quality than Europe (**Figure 2D**).

**Figure 2.**
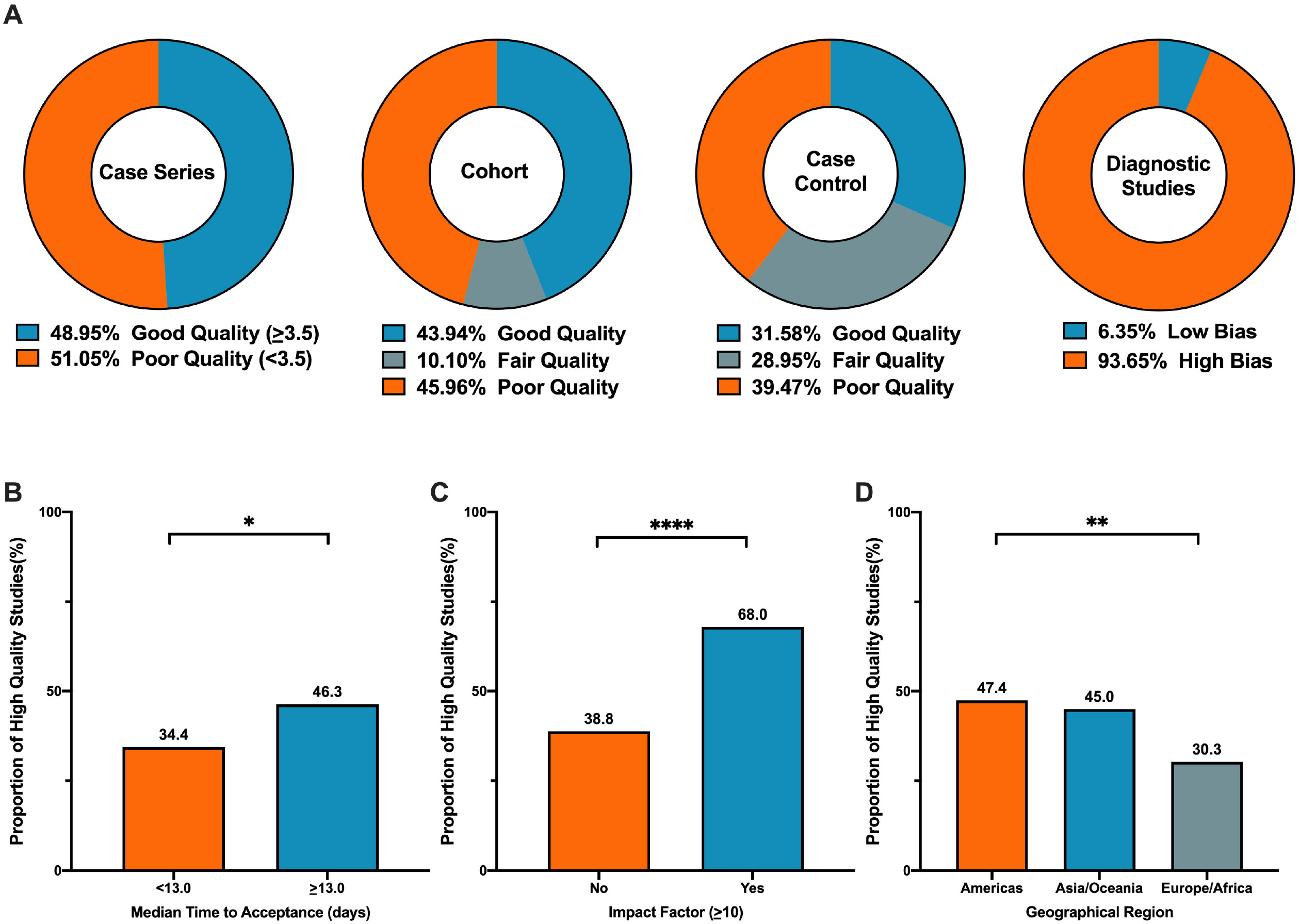
COVID-19 clinical literature quality assessment. **(A)** Overall good quality/low-bias studies represented less than half of identified works with 49.0% of case series, 43.9% of cohort, 31.6% of case-control, and 6.4% of diagnostic studies reporting good quality/low-bias. **(B)** Manuscripts with median time to acceptance <13.0 days had lower proportion of high study quality (34.4% vs. 46.3%, p=0.01). **(C)** Manuscripts with low impact factor (<10) had reduced proportion of high study quality (38.8% vs 68.0%, p<0.0001). **(D)** Studies originating in Americas or Asia/Oceania had higher study quality compared to that of Europe/Africa (47.4% vs. 45.0% vs. 30.3% for Americas, Asia/Oceania, Europe, respectively, p=0.01). Chi-Squares Test was conducted to evaluate differences in study quality by median time to acceptance, impact factor, and geographic region. P<0.05 was considered statistically significant.

### Methodological rigor differences in COVID-19 versus historical control

We matched 539 historical control articles to COVID-19 articles from the same journal in the previous year for a final analysis of 1078 articles (**Table 1**). Overall, the median time to acceptance was 13.0 (IQR, 5.0-25.0) days in COVID-19 articles vs. 110.0 (IQR, 71.0-156.0) days in control articles (**Figure 3A**, p<0.0001). Overall, methodological quality was lower in COVID-19 articles with 220 COVID-19 articles of high quality (41.0%) vs. 392 control articles (73.3%, **Figure 3B**, p<0.0001). High case series quality was observed 133 COVID-19 articles (48.0%) vs. 236 control articles (85.2%, **Figure 3C**, p<0.0001) with a difference in mean case series quality score (3.3 ± 1.1 vs. 4.3 ± 0.8, COVID-19 and control, respectively, p<0.0001). High cohort study quality was observed 76 COVID-19 articles (43.7%) vs. 129 control articles (74.1%, **Figure 3D**, p<0.0001) with a difference in mean cohort study quality score (5.8 ± 1.6 vs. 7.1 ± 1.0, COVID-19 and control, respectively, p<0.0001). High case-control study quality was observed 9 COVID-19 articles (28.1%) vs. 18 control articles (56.3%, **Figure 3E**, p=0.02) with a difference in mean case-control study quality score (5.4 ± 1.9 vs. 6.6 ± 1.0, COVID-19 and control, respectively, p=0.003). Finally, high diagnostic study quality was observed 12 COVID-19 articles (22.6%) vs. 24 control articles (45.3%, **Figure 3F**, p=0.01).

**Figure 3.**
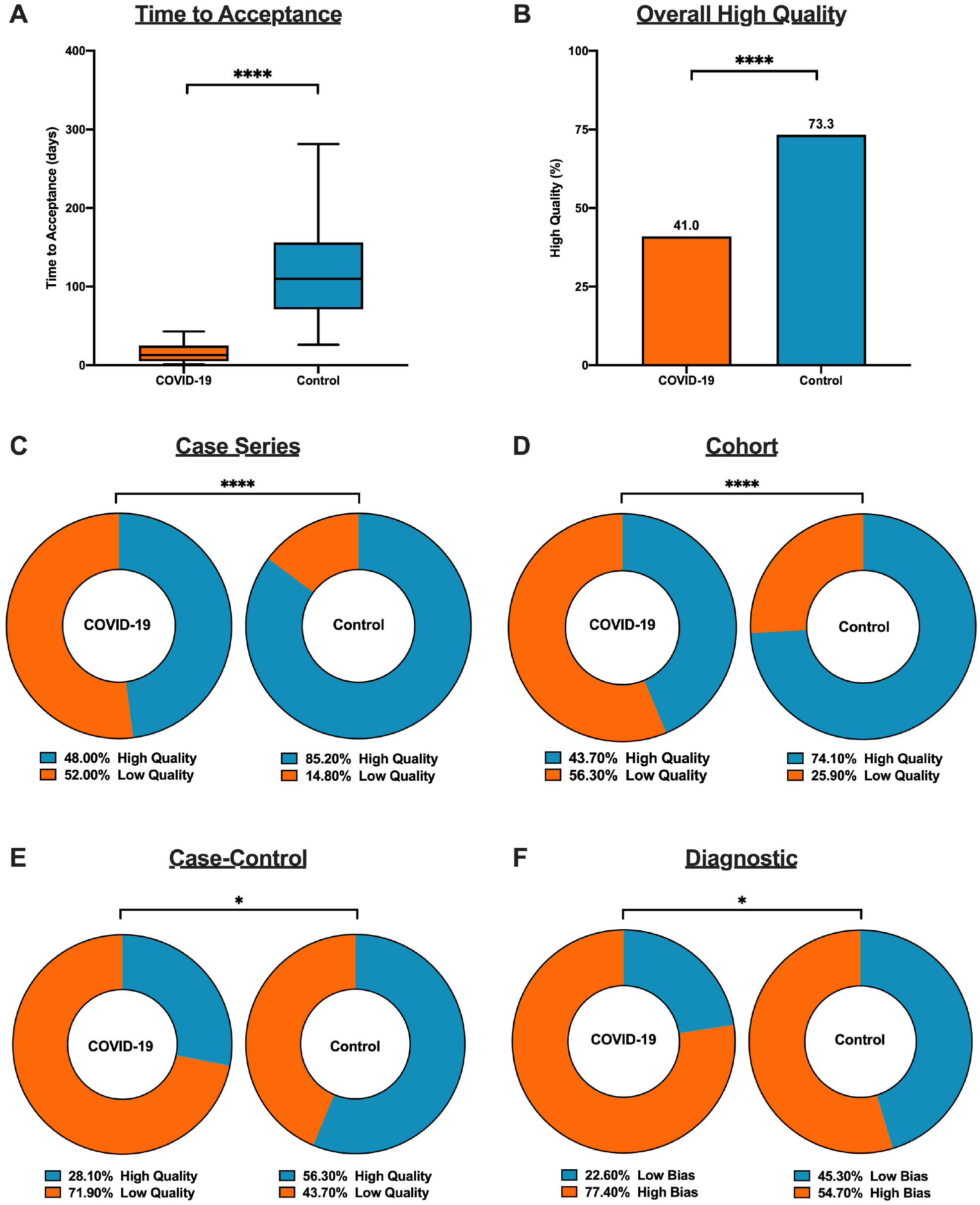
Differences in methodological quality in COVID-19 versus comparative article. **(A)** Median time to acceptance was reduced in COVID-19 articles compared to control articles (13.0 [IQR, 5.0-25.0] days vs. 110.0 [IQR, 71.0-156.0] days, p<0.0001). **(B)** Methodological quality was lower in COVID-19 articles compared to control articles (220 (41.0%) vs. 392 (73.3%), p<0.0001). **(C)** High quality case series studies was lower in COVID-19 articles compared to control articles (133 (48.0%) vs. 236 (85.2%), p<0.0001). **(D)** High quality cohort studies was lower in COVID-19 articles compared to control articles (76 (43.7%) vs. 129 (74.1%), p<0.0001). **(E)** High quality case-control studies was lower in COVID-19 articles compared to control articles (9 (28.1%) vs. 18 (56.3%), p=0.02). **(F)** Low bias in diagnostic studies was reduced in COVID-19 articles compared to control articles (12 (22.6%) vs. 24 (45.3%), p=0.01). Mann-Whitney U-test was conducted to evaluate differences in median time to acceptance between COVID-19 and control article. Differences in high study quality was evaluated by Chi-Squares Test. P<0.05 was considered statistically significant.

To determine the association of variables with higher study quality, we performed a logistic regression. In an unadjusted analysis, COVID-19 articles were associated with lower quality (odds ratio (**OR**), 0.25; 95% CI, 0.20 to 0.33, p<0.0001, **Table 2**). A multivariable logistic regression was generated including COVID-19 vs. control articles, median time to acceptance, geographical region, and high impact factor journal. Both increased time to acceptance and higher impact factor was associated with increased odds of higher study quality, whereas COVID-19 articles and publication originating from Europe/Africa were associated with reduced odds of higher study quality (**Table 2**).

**Table 2.**
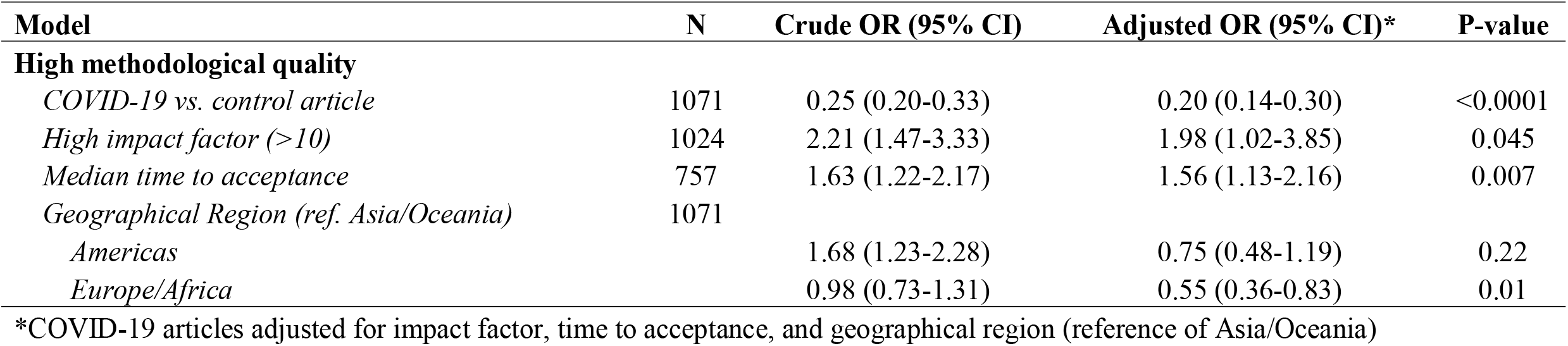
Association between high methodological quality and journal characteristics in COVID-19 clinical studies

## Discussion

In this systematic evaluation of methodological quality, COVID-19 clinical research was primarily observational in nature with modest quality. Not only were the study designs low in the hierarchy of scientific evidence, we also found that less than half of all studies met our prespecified threshold of quality. A longer peer-review process and publishing in higher impact journals was associated with improved rigor – albeit with modest improvements. In a case-control analysis with control articles identified from the same journal, we demonstrate reduced quality scores and a shorter time from submission to acceptance COVID-19 articles. Overall, the accelerated publication of COVID-19 research negatively affected the study quality compared to previously published comparative studies.

To our knowledge, this is the first time COVID-19 clinical study quality has been evaluated in a comparative study using a case-control design. Our research highlights major differences in study quality between COVID-19 and control articles driven by a combination of thorough peer review process as measured by increased time to publication in control articles and robust study design with questions which are pertinent for clinicians and patient management.^3 9-14^ Most importantly, our study highlights that a robust peer-review process and publication in leading medical journals with strict methodological guidelines and reporting are protective factors for publication of high-quality research.^15-17^

In the early stages of the COVID-19 pandemic, a hunger for data to inform clinical, social and economic decisions led to rapid dissemination and exponential publication of observational studies.^1 15^ This accelerated process allowed the understanding of natural history of COVID-19 and identification of tools to diagnose SARS-CoV-2; however, lower quality studies fundamentally risk patient safety, resource allocation, and future scientific research as these studies are based on flawed initial observations.^18^ Ultimately, poor evidence begets poor clinical decisions.^19^ This fundamentally risks undermining the public’s trust in science in this desperate time and has well been characterized through misleading information and high-profile retractions.^15 20-22^ Finally, early low quality studies can significantly decrease value of the scientific enterprise and increase waste of research funding replicating early poorly performed studies.^23^ Traditional peer-review process has been strained by the explosion of COVID-19 related articles with editorial boards revising their peer-review strategies to deal with an increase in submissions related to the field.^16 24^

Major breakthroughs in combating COVID-19 require properly designed studies which does not contribute to irreproducibility, resource wasting, and erroneous conclusions which may ultimately hinder progress.^5 18^ For example, hydroxychloroquine touted early in the pandemic has subsequently failed to be replicated in multiple observational studies and RCTs.^5 25-30^ One poorly designed study combined with rapid publication led to considerable investment of the scientific and medical community - akin to quinine being sold to the public as a miracle drug during the 1918 Spanish Influenza.^31 32^ Moreover, as of June 30, 2020, ClinicalTrials.gov lists an astonishing 230 COVID-19 trials with hydroxychloroquine/plaquenil, and a recent systematic review of observational studies and RCTs demonstrate no evidence of benefit nor harm with concerns of severe methodological flaws in the included studies.^33^

Our study is not without limitations. We evaluated the methodological quality of existing studies using established quality scores. While it is tempting to associate quality scores with reproducibility, it is not possible to ascertain the impact on the design and conduct of research nor results or conclusions in the identified reports.^34^ Second, our analysis includes early publications on COVID-19 and there is likely to be an improvement in quality of related studies and study design as the field matures and higher quality studies which take longer to design, conduct, and report are published. Accordingly, our findings are limited to the early body of research as it pertains to the pandemic and it is likely that over time research quality will improve.

## Conclusion

In summary, the early body of peer-reviewed COVID-19 literature was composed primarily of observational studies that underwent shorter time of evaluation and were of lower methodological quality than comparative studies in similar journals. Given the implications of these studies to medical decision making and government policy, greater effort to appropriately weigh the existing evidence in the context of emerging high-quality research is needed.

### What is already known on this topic

- The coronavirus disease 2019 (COVID-19) pandemic has devastated human society and an urgent need to diagnose and develop therapies has led to an explosion of COVID-19 literature
- COVID-19 publications have been rushed through peer-review due to the global emergency, but concerns exist regarding its methodological quality and false conclusions

### What this study adds

- Our study reveals majority of COVID-19 articles were of low hierarchy of evidence and less than half of COVID-19 articles were of high methodological quality, contributing to resource wasting, irreproducibility, and erroneous conclusions
- When compared to a case-control article, methodological rigor in COVID-19 articles were lower compared to control articles (41.0% vs. 73.3%, p<0.0001)
- Given the implications of these studies to medical decision making and government policy, greater effort to appropriately weigh the existing evidence in the context of emerging high-quality research is needed

## Data Availability

The statistical code and the entire dataset will be provided for transparency upon publication.

## Abbreviations

COVID-19: Coronavirus disease 2019
NOS: Newcastle-Ottawa Scale
OR: odds ratio
RCT: randomized controlled trial
SARS-CoV-2: Severe acute respiratory syndrome coronavirus 2

## Author contributions

**Study conception and design:** R. Jung, P. Di Santo, S. Visintini, F.D. Ramirez, T.Simard, and B. Hibbert

**Acquisition, analysis, or interpretation of data:** R. Jung, P. Di Santo, C. Clifford, G. Prosperi- Porta, S. Parlow, S. Skanes, A. Hung, F.D. Ramirez, T. Simard, and B. Hibbert

**Drafting of the manuscript:** R. Jung, P. Di Santo, F.D. Ramirez, T. Simard, and B. Hibbert

**Critical revision of manuscript**: All authors

**Statistical analysis:** R. Jung, P. Di Santo and B. Hibbert

**Supervision:** B. Hibbert

**Guarantors of the study:** R. Jung, P. Di Santo, and B. Hibbert.

The corresponding author attests that all listed authors meet authorship criteria and no others meeting the criteria have been omitted.

## Funding/Support

This study received no specific funding or grant from any agency in the public, commercial, or not-for-profit sectors. R. Jung was supported by the Vanier CIHR Canada Graduate Scholarship. F.D. Ramirez was supported by a CIHR Banting Postdoctoral Fellowship and a Royal College of Physicians and Surgeons of Canada Detweiler Travelling Fellowship. The funder/sponsor(s) had no role in design and conduct of the study, collection, analysis, and interpretation of the data.

## Competing interest

All authors have completed the ICMJE <u>Unified Competing Interest form</u> (available on request from the corresponding author) and declare: no support from any organisation for the submitted work; no financial relationships with any organisations that might have an interest in the submitted work in the previous three years, no other relationships or activities that could appear to have influenced the submitted work. B. Hibbert reports funding as a clinical trial investigator from Abbott, Boston Scientific, and Edwards Lifesciences outside of the submitted work.

## Ethical approval

Not required.

## Transparency declaration

The corresponding author affirms that the manuscript is an honest, accurate, and transparent account of the study being reported; that no important aspects of the study have been omitted; and that any discrepancies from the study as planned (and, if relevant, registered) have been explained.

## Data sharing

The statistical code and the entire dataset will be provided for transparency.

## Dissemination to participants and related patient and public communities

We plan to disseminate the findings of the study in collaboration with our media relations division through social media, press release, and journal publication.

## Notes

### Competing Interest Statement

All authors have completed the ICMJE Unified Competing Interest form (available on request from the corresponding author) and declare: no support from any organisation for the submitted work; no financial relationships with any organisations that might have an interest in the submitted work in the previous three years, no other relationships or activities that could appear to have influenced the submitted work. B. Hibbert reports funding as a clinical trial investigator from Abbott, Boston Scientific, and Edwards Lifesciences outside of the submitted work.

